# A Two Parameter Closed Form Solution to Kermack and McKendrick’s Integro-Differential Equations

**DOI:** 10.1101/2023.11.13.23298463

**Authors:** Ted Duclos, Tom Reichert

## Abstract

A closed form solution of the full integro-differential equations in the 1927 paper by Kermack and McKendrick (K&M), called herein the KMES, is presented and verified. The solution arises from the use of the time-based weighted averages of K&M’s rate parameters to transform their equations into equivalent, solvable differential equations. Then, by using a network formulation, we find functional forms of the time varying parameters of these new equations, and derive the total case count as the integral of the product of the probability rate of transmission and the number of infectious contacts.

Analytical expressions for managing an epidemic, the real-time effective reproduction number, time to peak in new infections, and the final epidemic size, flow directly from the solution. Notably, although the KMES is derived from K&M’s equations, the expressions for time-to-peak and final size are antithetical to currently accepted epidemic concepts

The KMES has only two parameters: the basic reproduction number, and the probability rate of disease transmission, both of which can be readily derived from early epidemic data. Using early COVID-19 pandemic data from six different countries to estimate these two parameters, the KMES accurately projects case data from pandemic in the following 30 & 60-day periods with R^2^ values >0.93 and 0.78 respectively. In addition, a projection of a typical individual’s contagiousness, derived from the KMES, closely approximates the time course of viral shedding measured in infected persons.

## Introduction

Modern epidemiological modeling is rooted in the seminal paper of Kermack and McKendrick (K&M, 1927). Since its publication, well over 10,000 authors have referenced this paper as a foundational starting point. However, as Diekmann points out in his insightful essay (2022, pg. 8), “…an incessant community-enforced misconception is that the paper is just about the very special case, the S-I-R (Susceptible, Infected, Recovered) model.” In the essay, he also states his suspicion that, despite the numerous references, the citing authors rarely read the 1927 paper. His concluding remarks decry the situation and request that researchers carefully study the 1927 paper, a “…true gem…in which tremendous wisdom lies hidden.” (Diekmann 2022 pg. 9).

To Diekmann’s last point, we offer here that, a close reading of K&M’s 1927 paper reveals that the constants in the SIR model, typically denoted as *β* and *γ*, originate, in fact, from the assumption that the weighted averages of K&M’s parameters, *φ*(*θ*) and *ψ*(*θ*) (where *θ* denotes the time-since-infection), can be constants. Yet, despite purposeful searching, *β* and *γ* do not appear to ever have been referenced as weighted averages, and, consequently, no analyses can be found to confirm that the behavior of the original equations is retained when assuming the weighted averages are constant. This is an unfortunate oversight, since, as we show in this manuscript, the SIR model exhibits behavior not found in the solution to K&M’s full equations.

The assumption of constant weighted averages imposes the implicit assumption that the population is well mixed, a supposition Brauer (2008, pg. 27) labelled as “unrealistically simple”. In turn, the well-mixed assumption requires a priori knowledge of a third SIR parameter, the initial size of the susceptible population (commonly labelled, *S*(0)).

This presents health officials with a paradox: Both *β* and *S*(0) are proxies of population interaction, yet, in the SIR model, when the population interaction is increased by increasing one or the other, the time to the peak in infections either decreases (increasing *β*) or increases (increasing *S*(0)). Since Brauer (2008) also points out that *βS*(0) is the rate of the contacts that are sufficient to transmit an infection, the question, central to public health officials, becomes: Does a decrease in the contact rate move the peak time further out or bring it closer in time?

A solution to K&M’s full integro-differential equations could answer this question as well as unlock further wisdom, but the widespread use of SIR approximations (e.g., Breda et al 2021, Diekmann 2022 and Brauer 2008) and the recent publication of a discrete time model of the integro-differential equations (Diekmann 2021), imply that such a solution remains elusive. Absent a solution, researchers have made prodigious efforts to extract wisdom from the SIR approximations; and have produced the well-known formulas for the final size of epidemics, the notion of “flatten the curve” (DiLauro et al 2021), and various other refinements (e.g., Brauer 2008, Breda et al 2021, Hethcote 2000).

One alternative approach, network-based stochastic models (e.g., Newman 2002, Diekmann et al 1998, Youssef and Scoglio 2011, and Zonta and Levitt 2022) assumes that epidemic dynamics can be projected by summing the collective probabilities that individuals will be infected by a limited number of contacts; thereby avoiding the well-mixed assumption. These widely investigated approaches have produced estimations of outbreak threshold conditions and the final size of epidemics. However, they do not yield the type of analytical expressions which make closed-form solutions so appealing and useful.

The principal difficulty in finding a solution to K&M’s equations has been the challenging task of finding the functional forms for *φ*(*θ*) and *ψ*(*θ*). The simplest tactic to address this difficulty, assuming that the weighted averages of the parameters are constant, results in the SIR model, but does not yield solvable equations. By assuming that the weighted averages could vary in time in a fixed proportionality, Kroger and Schlickeiser (2021) did solve a special case of the SIR model equations, however, due to the proportionality constraint, the solution is not a general result.

In this manuscript, we find a general solution for K&M’s equations by allowing time varying weighted averages and employing a vision of an epidemic as a network. The network concept generates functional forms for the time varying weighted averages and reduces K&M’s equations to an equivalent system of solvable differential equations. This approach, apparently unexplored, yields a solution dependent on only two parameters and does not require a priori knowledge of *φ*(*θ*), *ψ*(*θ*), or the total population size.

Our solution closely follows Diekmann’s (2022) insight that the infection incidences should represent “…the probability per unit of time of (a) susceptible (person) becoming infected…times the susceptible subpopulation”. Since this also requires that there be contact between susceptible and infectious persons for incidences to occur, the solution for the time course of the total incidences should be the integral, over time, of the product of the probability rate of infection transmission and the number of infectious contacts with susceptible people; and, indeed, this is the result that we produce.

Inspired by Diekmann’s comments, we offer here a path to a closed form solution which we call the KMES. Our approach avoids the difficulties associated with K&M’s formulation, frees the analysis from the unrealistically simple assumption of a well-mixed population and reveals fundamental attributes of the K&M equations:

1. In Section 1 we solve the K&M integro-differential equations and demonstrate the veracity of the closed form both graphically and analytically. We then derive analytical expressions of real-world epidemic dynamics that arise directly from the solution.
2. In Section 2, using data compiled from six countries during the COVID-19 pandemic, we illustrate that the solution accurately projects the course of the COVID-19 pandemic in six sampled countries and provides an estimate of the related quantity, individual contagiousness.
3. In Supplements 2 and 3, we derive and demonstrate the use of additional expressions which can aid public health officials to diagnose, control, and even end an epidemic.

## Section 1: Solution to the Kermack and McKendrick Equations

We first convert K&M’s integro-differential equations into simpler, SIR differential equations with variable coefficients. We then use a network description of an epidemic to derive analytic expressions for the coefficients; and with these unveil the solution.

### Kermack and McKendrick’s Integro-Differential Equations

The terms used in K&M’s model are: *N*_*p*_, the total population; *A*_*p*_ the area that contains *N*_*p*_, and *S*(*t*), the portion of *N*_*p*_ that has not yet become infected. The ever-infected population is *N*(*t, θ*), the persons who are currently or have previously been infected (the cases); where *θ* is the time since infection of people within *N*(*t, θ*); and *N*(*t, θ*) is subdivided between *I*(*t, θ*), the individuals who are currently infectious and R(*t, θ*) the recovery of individuals within *N*(*t, θ*). We call *S*(*t*), *N*(*t, θ*), *I*(*t, θ*), and R(*t, θ*) the “population variables”, and call out that a population variable integrated over *θ* from 0 → *t* is a function of time alone (e.g.,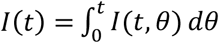). We note further that K&M defined *φ*(*θ*) as the probability rate of new infection transmission per susceptible density and infectiousness; and *ψ*(*θ*) as the probability rate of recovery per infectiousness.

Given these definitions, *N*(*t*) = *I*(*t*) + R(*t*), and S(0) = *N*_*p*_. We further assume that *N*_*P*_ is a constant, *I*(0) = *N*(0) = 1, R(0) = 0, and that, once infected, people cannot become reinfected. Also, since *θ* has the units of time, Δ*θ* = Δ*t*, and *θ* has values from 0 to t.

K&M derived their equations by imagining *N*(*t*) as the sum, at time t, of incremental subpopulations, 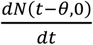, which we shall here call “*θ*-groups”, infected at time *t* − *θ*. They tracked the remaining infectiousness of each *θ*-group through both time and *θ* using an infectiousness probability function, 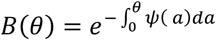, and constructed their equations by tracking the recovered and still infected portions of *N*(*t*). K&M’s resulting integro-differential equations are:

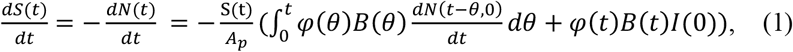

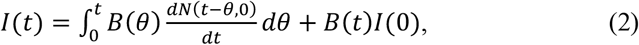

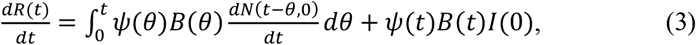

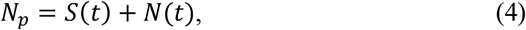

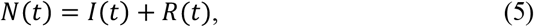

Although K&M (1927) wrote their equations in terms of subpopulation densities, we purposefully chose, without altering their result, to express them in terms of subpopulation totals.

Equations 1 through 5 can be reduced to a system of differential equations by first noting that 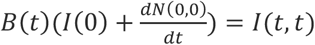, thus, 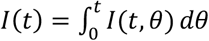 in Equation 2, and Equations 1 through 3 can be equivalently written as,

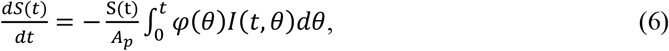

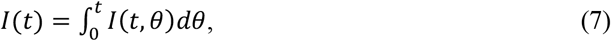

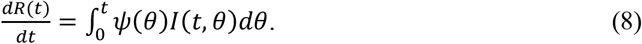

Dividing Equations 6 and 8 by Equation 7, we find the time-based weighted averages of *φ*(*θ*) and *ψ*(*θ*),

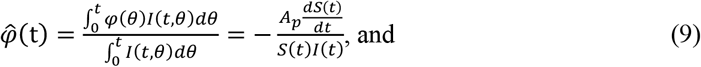

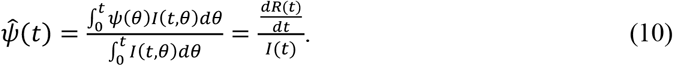

We then use these weighted averages to convert K&M’s equations into the following equivalent system:

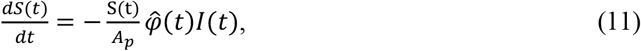

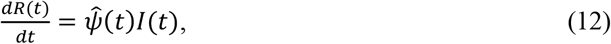

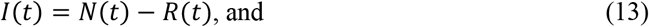

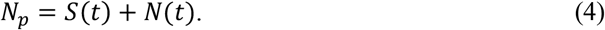

If we set 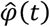 and 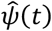 equal to the constants *β* and *γ*, then this system of equations simplifies to the SIR model with a well-mixed population. Thus, as stated in the introduction, the constants *β* and *γ* in the SIR model are the weighted averages of *φ*(*θ*) and *ψ*(*θ*). These and a third parameter, *S*(0) are needed to solve the Equations in this form.

##### Sidebar

###### Using the Weighted Averages

Here we explain the relationships between *φ*(*θ*) and *ψ*(*θ*) and their weighted averages.

The time since infection, *θ*, has the units of time, has values on the interval 0 < *θ* ≤ *t*, and can also be expressed in terms of time. Therefore, the discrete form of Equation 8 when using 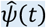 or *ψ*(*θ*) is:

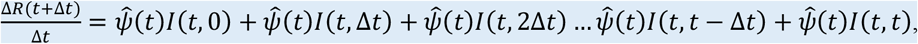

or

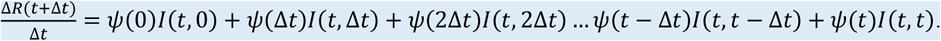

From this, we can see that although the value of 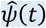 does not necessarily equal any of the individual values of *ψ*(*θ*), it does become fixed as the single value at time t which, when multiplied by the individual values of *I*(*t, θ*) results in the same sum as when the multiplication is performed using *ψ*(*θ*).

We do not need to know the individual values of *ψ*(*θ*) to calculate 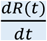. We only need a method to calculate 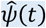 and to be able to sum the *I*(*t, θ*) values. We also do not need to know the individual values of *I*(*t, θ*); we merely need to constrain the total of the *I*(*t, θ*) values to sum to the proper value of *I*(*t*). These comments also apply to the substitution of 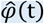 for *φ*(*θ*) in Equation 6 to obtain Equation 11.

###### The *B* Function

As formulated by K&M (1927), the function *B*(*θ*), derives from using *ψ*(*θ*) to calculate the infectiousness of each *θ*-group in prior time intervals all the way back to the time of infection for each *θ*-group; *B*(*θ*) is, in discrete form,

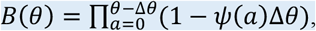

which means 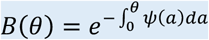 in the limit as Δ*θ* → 0.

The analog to this expression, found by the same method as was used to find *B*(*θ*), but using 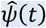 is,

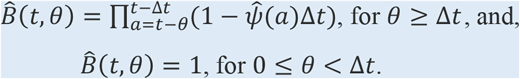

Consequently, 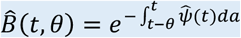 in the limit as Δ*t* → 0.

The interval between the integral limits in the expression for 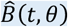 is *θ*, the same length as the interval in *B*(*θ*). This ensures that the proper values of 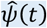 are used in the calculation of the infectiousness of each *θ*-group.

Because of how they were calculated, *B*(*θ*), and 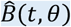 both determine the total residual infectiousness at time t of the previously and newly infected people, and therefore,

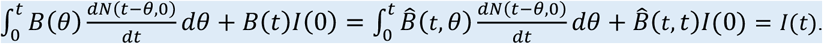

A detailed derivation of this result is provided in Supplement 1.

### The Epidemic as a Network

To solve Equations 11, 12, 13 and 4, we must first find expressions for 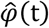 and 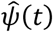; and to accomplish this we turn to a network depiction of the epidemic.

We begin by envisioning the population affected by the epidemic as a network represented by a connected graph with *N*_*P*_ persons/vertices. The network edges are the contacts of the *N*_*P*_ persons with an infection transmission probability of one if the node remains sufficiently contagious. Since the transmission rate is finite, an infected person could recover before transmitting the infection along all its edges, and each edge might possibly not pass on the infection.

We assume that all contacts other than the edges have an infection transmission probability of zero, hence, the mean number of the edges per person is the self-weighted mean degree of this network minus 1; and, by definition, this is also the basic reproduction number, R_0_. That is to say, if an epidemic progresses within *N*_*P*_, this conceptualization of *R*_0_ translates as each member of *N*(*t*) being associated with a group of contacts of average size *R*_0_. We label these groups “N-groups”; and note that persons within *N*(*t*) are not members of their own N-groups.

This notion of *R*_0_ is consonant with a conventional concept of *R*_0_, which states that, “*R*_0_ = expected number of secondary cases per primary case in a ‘virgin’ population.” (Diekman et al 2013 pg. 4). This is represented mathematically as *R*_0_ = *pc*(*t*_1_ − *t*_2_) (Diekman et al 2013, pg. 4); where *p* is the probability a contact will become infected, *c* is the expected contact rate, and *t*_1_ − *t*_2_ is the period when *p* is non-zero.

In our conception, since *R*_0_ is the number of contacts in the “virgin” population that will become infected, *p* = 1 during the period *t*_1_ − *t*_2_; and, consequently, *R*_0_ = *c*(*t*_1_ − *t*_2_). We do not need to specify either the period *t*_1_ − *t*_2_ or the contact rate *c* hidden within *R*_0_ to derive our solution; however, later in our analysis, we apply a probability rate of transmission to capture the dynamics of infection transmission.

Of course, not everyone who is infected can transmit the disease. Therefore, we distinguish three subgroups of infected people. These are:

1. <u>Latently Infectious</u>: People who have been infected but are not yet shedding virus.
2. <u>Actively Infectious:</u> People who are shedding the virus and <u>are in contact</u> with susceptible people.
3. People who may be shedding the virus but have <u>no contact</u> with susceptible people.

These are effectively and mathematically part of the recovered group, R(*t*).

Persons in groups 1 and 2 constitute *I*(*t*) and are collectively known as infectious. Persons in group 2 are infectious and contagious, and persons in group 3 are contagious, but not infectious. All three groups, together with the previously infected, but now recovered group, comprise the ever-infected population, *N*(*t*).

We assume in our construction that infectors remain in durable, potentially infectious contact with the people they have infected. Consequently, the infectiousness of the infectors, but not their degree within the network, <u>diminishes</u> with each new infection they cause. In symmetry, the extent to which they are unable to infect others, their recovery increases. We also assume in our analysis that *R*_0_ stays constant, no two N-groups are identical, and, for simplicity, that the disease does not have a latent period (i.e., *I*(*t*) in this analysis is solely people in group 2).

### The Solution

In contrast to K&M’s *θ*-group formulation, the N-group perspective enables us to determine expressions for 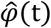 and 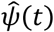 in terms of time, a rate of infection, and *R*_0_.

The totality of the contacts in the N-groups can be divided into two time-varying portions: R_*eff*_(*t*), the average number of contacts within each N-group that remain susceptible; and R_*N*_(*t*), the average number of previously infected contacts. Given that the susceptible persons within the N-groups are, by definition, in contact with an infectious person, 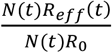 is the probability that a member of *N*(*t*) is infectious (i.e., 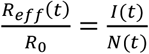). Thus,

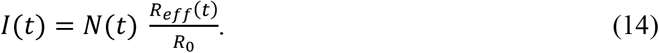

Since the product of 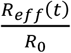 and *N*(*t*) yields the total residual infectiousness of the *N*-groups, Equation 14 is the N-group analog to Equation 2.

If we define 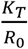 as the probability rate of transmission between susceptible and infectious contacts, assume it to be constant across the population, and assume *K*_*T*_ is a quantity associated with the disease, then,

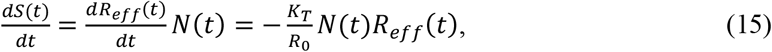

Analogous to Equation 14, since the product of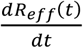 and *N*(*t*) yields the total change in the susceptible population, Equation 15 is the N-group equivalent of Equation 1.

We know that 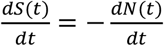, therefore, we can use this relationship and Equation 14 to restate Equation 15 as,

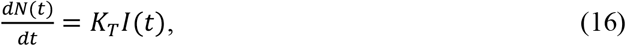

which, based on Equation 11 leads us to an expression for 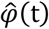,

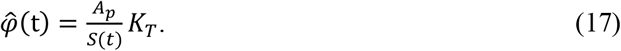

We observe in Equation 17 that, in contrast to the SIR equations, the N-group perspective does not need the value of *S*(0) a priori to evaluate 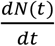.

To find an expression for 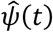, we begin with the N-group formulation of the recoveries,

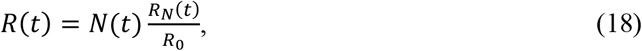

and differentiate to find,

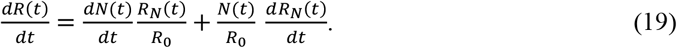

Then, since R_*eff*_(0) = *R*_0_, integrating and exponentiating Equation 15 produces,

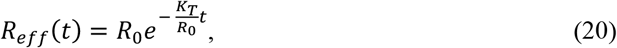

Applying Equation 14 to Equation 20, and defining *F*_*i*_(*t*) as the fraction of *N*(*t*) that is infectious,

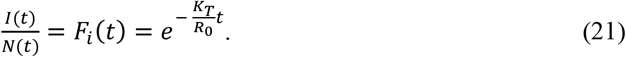

We remark that, R_*N*_(*t*) = *R*_0_ − R_*eff*_(*t*), and by Equation 20,

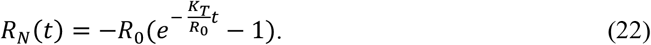

Differentiating Equation 22, we obtain,

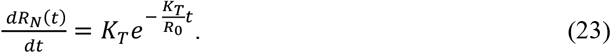

Using Equations 21, 22, 23, and 16, Equation 19 becomes,

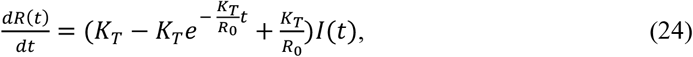

and, by Equation 12 and 24, we find the expression for 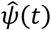,

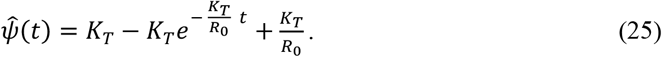

Having found 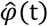 and 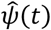 we can now write K&M’s equations as,

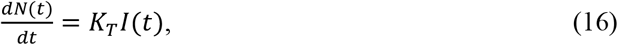

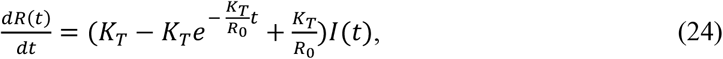

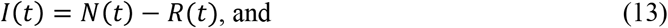

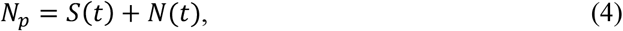

which is a system of equations equivalent to Equations 1 through 5.

Though the analysis should be clear, we offer the following comments to help a reader intuit how the N-group perspective transforms K&M’s equations into this system.

First, the notion that infectiousness is reflective of contact with susceptible persons, embodied in the change in perspective from *θ*-groups to N-groups and typified by Equation 15, subsumes *S*(*t*) into *K*_*T*_. A constant rate of transmission between infectious people and their eventual recipients means that 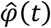 and *S*(*t*) are inversely proportional, a result congruent with their units.

Second, the N-group perspective enables us to get a sense of the incremental recovery function in Equation 24. We first note that 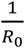 is the portion of the new infections, *K*_*T*_ *I*(*t*), that diminish the infectiousness of the infectors as they incrementally infect their N-groups. Therefore, since *K*_*T*_*I*(*t*) is also the incremental increase in *N*(*t*), and the equivalent terms 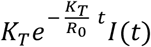 and 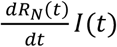 are each the portion of the change in *I*(*t*) that is newly infected, then the difference, 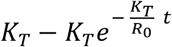, equates to the portion of the existing infectiousness that has just recovered.

The solution to Equations 16, 24, 13, and 4 is,

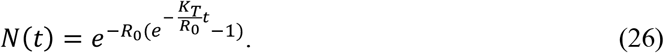

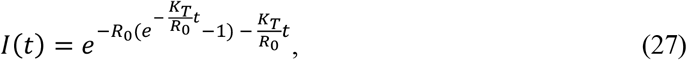

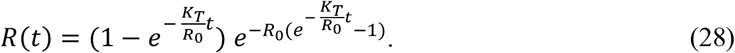

We call Equations 26 through 28 the KMES (**<u>K</u>**ermack and **<u>M</u>**cKendrick **<u>E</u>**quation **<u>S</u>**olution) and maintain that they are a solution to Equations 1 through 5. We provide graphical and analytical proof supporting this claim in the next two subsections.

The logic of the KMES emerges if one first observes that the term 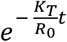 is the probability that any member of *N*(*t*) is infectious or that a member of the N-groups is susceptible. Therefore, the expressions for *I*(*t*) and R(*t*) are merely the product of *N*(*t*) times 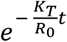 and 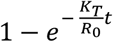 respectively. Additionally, as anticipated by Diekman’s statement, the expression for *N*(*t*) itself is, indeed, the integral, up to the current time t, of the product of the probability rate of transmission 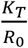, and the number of infectious contacts *I*(*t*)*R*_0_.

### Graphical proof

To show, graphically, that the KMES projects the same time course of *N*(*t*), *I*(*t*), and R(*t*) as K&M’s integro-differential equations, we use Equations 16, 24, 13, and 4, since this system of equations is equivalent to Equations 1 through 5. To this end, we used the parameters listed in Table 1 in a fourth order Runga-Kutta algorithm to simulate Equations 16, 24, and 13, and plot these alongside the KMES in Figure 1. This figure illustrates that the KMES replicates the simulation with a vanishingly small error, proving that the KMES solves K&M’s equations.

**Table 1.** Values of the parameters used to create Figure 1.

|  |  |  |  |  |  |  |  |  |  |  |
| --- | --- | --- | --- | --- | --- | --- | --- | --- | --- | --- |
| Condition A | Time step= | 0.1 | KT= | 0.26 | Ro= | 10 | I(0)= | 1 | Np= | 40000000 |
| Condition B | Time step= | 0.1 | KT= | 0.26 | Ro= | 11 | I(0)= | 1 | Np= | 40000000 |
| Condition C | Time step= | 0.1 | KT= | 0.26 | Ro= | 12 | I(0)= | 1 | Np= | 40000000 |

**Figure 1.**
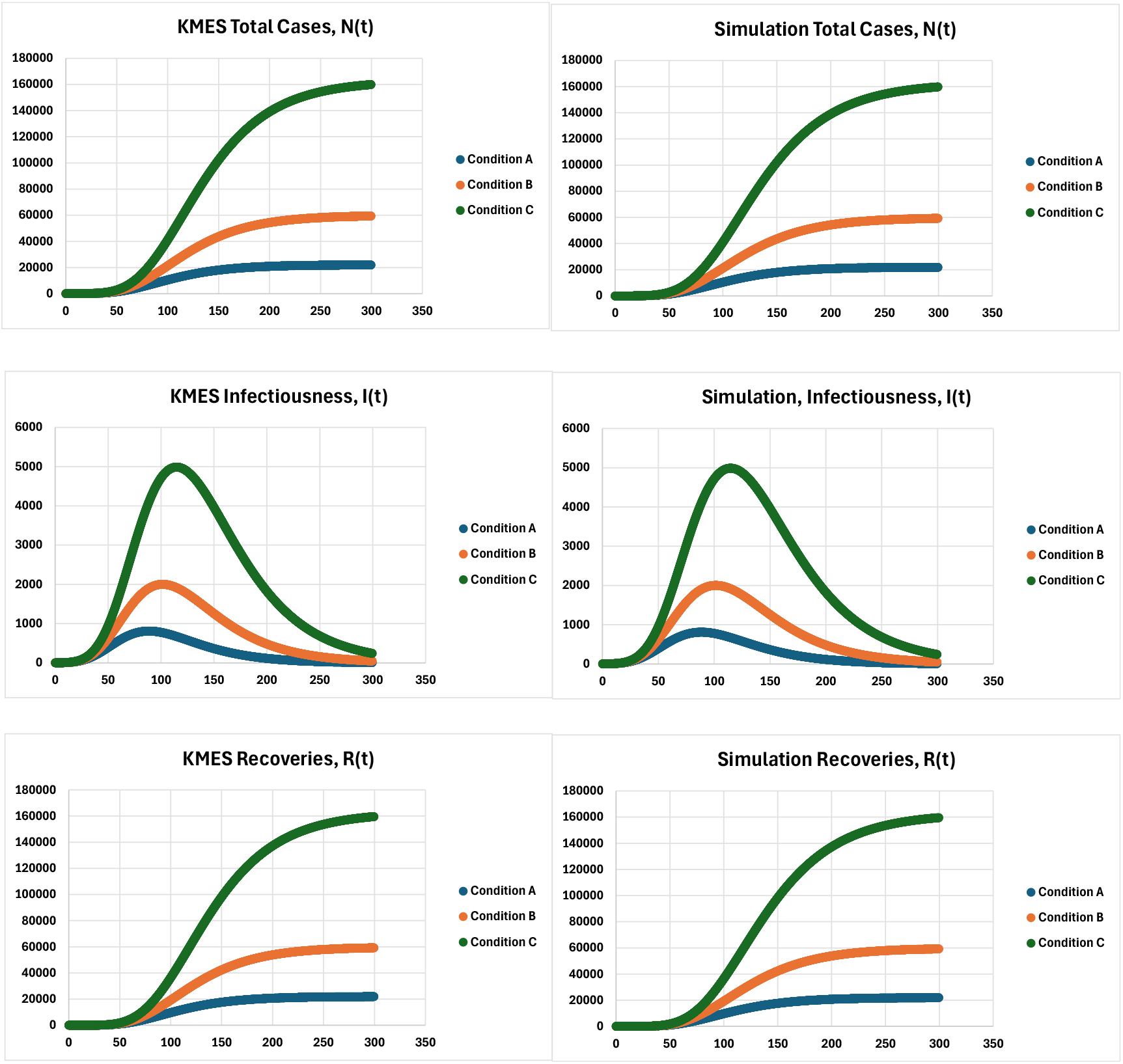
Calculations of *N*(*t*), *I*(*t*), and *R*(*t*) using the KMES (Left Side; Equations 26-28), and Simulation of Equations 16, 24, and 13 (Right Side). The parameter values found in Table 1 were used to generate the plots. The projections appear in separate plots because the Mean Absolute Percent Error between them is less than 5E-7, and if they were plotted together, then the top plot would completely obscure the underlying plot.

### Analytical proof

As a check on the graphical proof, we can insert the KMES into the *θ*-based Equations 1-5 and show analytically that it solves them. We begin by solving Equation 2, a task that uses the outcome explained in the Sidebar and Supplement 1 that,

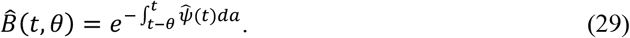

and that 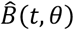 can be substituted for *B*(*θ*) in Equations 1 through 3 without affecting the values of the equations.

Substituting the right side of Equation 25 into Equation 29, we obtain,

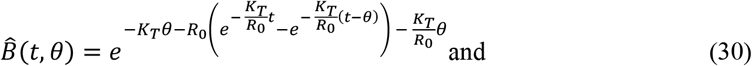

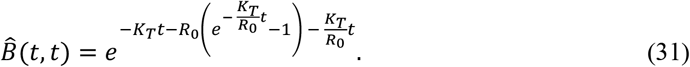

Then, by differentiating both sides of Equation 26 and using *t* − *θ* for time, we find,

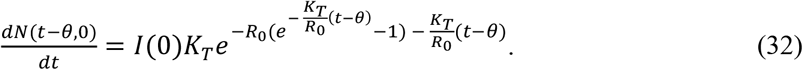

Substituting these three expressions into Equation 2, we have,

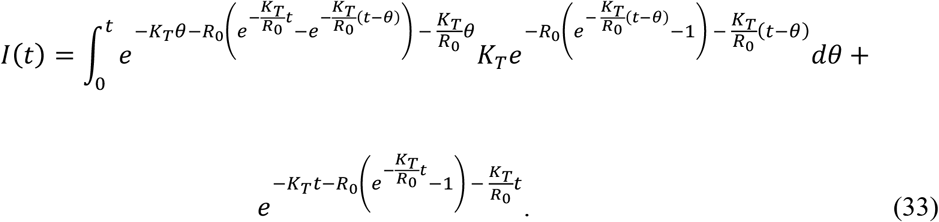

Evaluating the integral in Equation 33 and simplifying, we obtain,

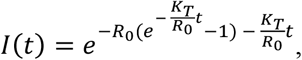

which thereby proves that Equation 27 is the solution for *I*(*t*) in Equation 2.

By using the relationship 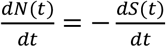 and the weighted averages in Equations 9 and 10, we can rewrite Equations 1 and 3 as,

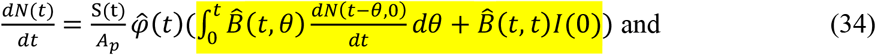

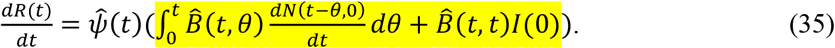

By Equation 2, the expressions highlighted on the right-hand side of both Equation 34 and 35 equal *I*(*t*). Substituting the proven expression for *I*(*t*) along with Equations 9 and 10 into Equations 34 and 37, we find,

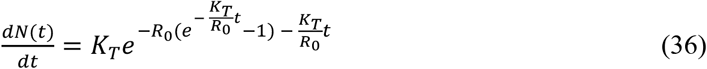

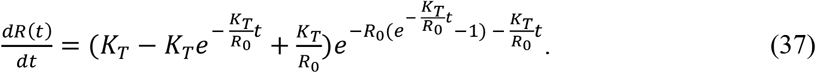

Both equations can be integrated to find the remainder of the solution,

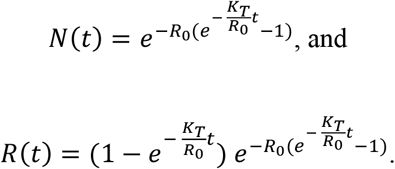

Thus, the KMES is a closed form solution to K&M’s integro-differential equations.

### Expressions Describing an Epidemic

Because it is a closed form solution, expressions useful in epidemic description and management emerge from the KMES. We illustrate a few of these here.

### Epidemic Direction

Using the expression for *R*_*eff*_(*t*), Equation 20, to rewrite the derivative of Equation 27 produces,

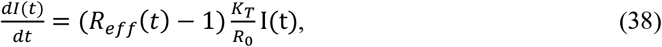

Therefore, if *R*_*eff*_(*t*) > 1, the epidemic is growing, and reciprocally, if *R*_*eff*_(*t*) ≤ 1, the epidemic is declining. As demonstrated in Section 2, *K*_*T*_ and *R*_0_ can be evaluated using early epidemic data. Therefore, Equation 20 can assist officials in making public health decisions.

### Time to Peak New Infections and Peak Size

Setting *R*_*Eff*_ (*t*) = 1 in Equation 20 yields the time when both *I*(*t*) and 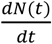 peak,

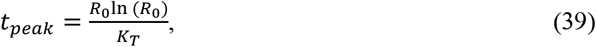

Equation 39 directly answers the question, raised in the introduction, whether the peak in infections comes sooner or later if the social interactions are decreased: The peak comes sooner. Since this is the converse of the SIR model phenomenon known as “flatten the curve”, Equation 39 strongly suggests that this historical notion should be abandoned.

Usefully, Equation 39, when substituted into Equation 27, generates an equation for projecting the peak value of *I*(*t*),

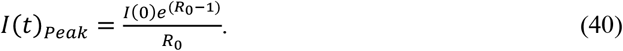

### Final Size

In the limit as *t* → ∞, Equation 13 simplifies to an expression for the final size,

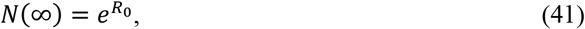

Therefore, if *R*_0_ < ln (*N*_*P*_), then the entire population will not become infected. However, if *R*_0_ remains large enough for long enough (i.e., *R*_0_ ≥ ln (*N*_*P*_)), the KMES projects that *N*(*t*) <u>can</u> equal *N*_*P*_ and herd immunity is not guaranteed.

This result differs from K&M’s (1927) conclusion that the entire susceptible population cannot be infected; an outcome that has achieved a position of prominence within the epidemiological modelling community. Nevertheless, since Equation 41 derives from the KMES, and the KMES solves Equations 1 through 5, we must conclude that the K&M integro-differential equations do, in fact, allow for an entire population to become infected.

K&M’s (1927) conclusion rested upon their assumption that the expression,

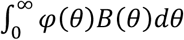, remains finite for all time. This assumption, however, is mathematically unsupportable, because in Equation 1, if 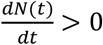 (i.e. new infections are occurring) and *t* ≠ ∞, then as *S*(*t*) goes to zero, both 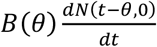 and *B*(*t*)*I*(0) remain finite and greater than zero. Therefore, consistent with Equation 17, *φ*(*θ*) must trend to infinity and 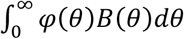 can, indeed, become unbounded.

Since *φ*(*θ*) trends to infinity as *S*(*t*) goes to zero, its weighted average must also go to infinity. Therefore, we cannot assume 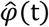 is a constant, as the SIR model does, and also expect that model to retain key aspects of either K&M’s original equations or their solution.

## Section 2: Projections of Pandemic Data Using the KMES

If we differentiate both sides of Equation 26, divide by *N*(*t*), rearrange using *F*_*i*_, and take the natural log of both sides, we find an expression useful for making projections,

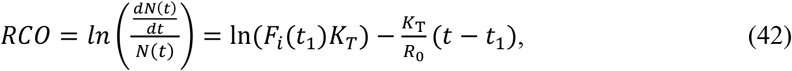

where *t*_1_ < *t*. We have labelled this quantity the “Rate of Change Operator” (RCO) because it is a measure of the rate of change of *N*(*t*) per person within *N*(*t*). With the RCO, we can extract the values of *F* _*i*_ (*t*_1_)*K* _*T*_ and 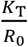 from case data; and, since Equation 26 may be transformed to,

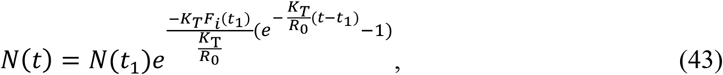

we can use Equations 42 and 43 to project the total and new cases during an epidemic.

To demonstrate this process, we applied Equation 42 to data recorded in six different countries during the initial stages of the COVID-19 pandemic (Roser, et al. 2021), and created the plots in Figure 2. These plots show that the RCO trends were linear both before and shortly after the date of the imposition of containment actions (arrows in the plots), indicating that 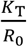 remained constant in those periods. By fitting Equation 42 to short, nine data point portions early in the straight segments of each country’s RCO time series, we then estimated values of both 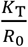 and *F*_*i*_(*t*_1_)*K*_*T*_ for these countries and tabulated them in Table 2.

**Table 2.** Social containment parameters used to model total cases and new daily cases of infection for different countries (Roser et al 2021).

| | $\frac{K_T}{R_0}$ | $\ln(F_i(t_1)K_T)$ | $N(t_1)$ | Date range for RCO fit |
| --- | --- | --- | --- | --- |
| **South Korea** | 0.24 | -1.58 | 3,736 | March 1–March 9 |
| **USA** | 0.076 | -1.39 | 46,136 | March 23–March 31 |
| **Sweden** | 0.036 | -2.47 | 5,320 | April 1–April 9 |
| **Italy** | 0.080 | -1.93 | 31,506 | March 17–March 25 |
| **Spain** | 0.09 | -2.11 | 65,719 | March 27–April 4 |
| **New Zealand** | 0.20 | -0.89 | 155 | March 24–April 1 |

**Figure 2.**
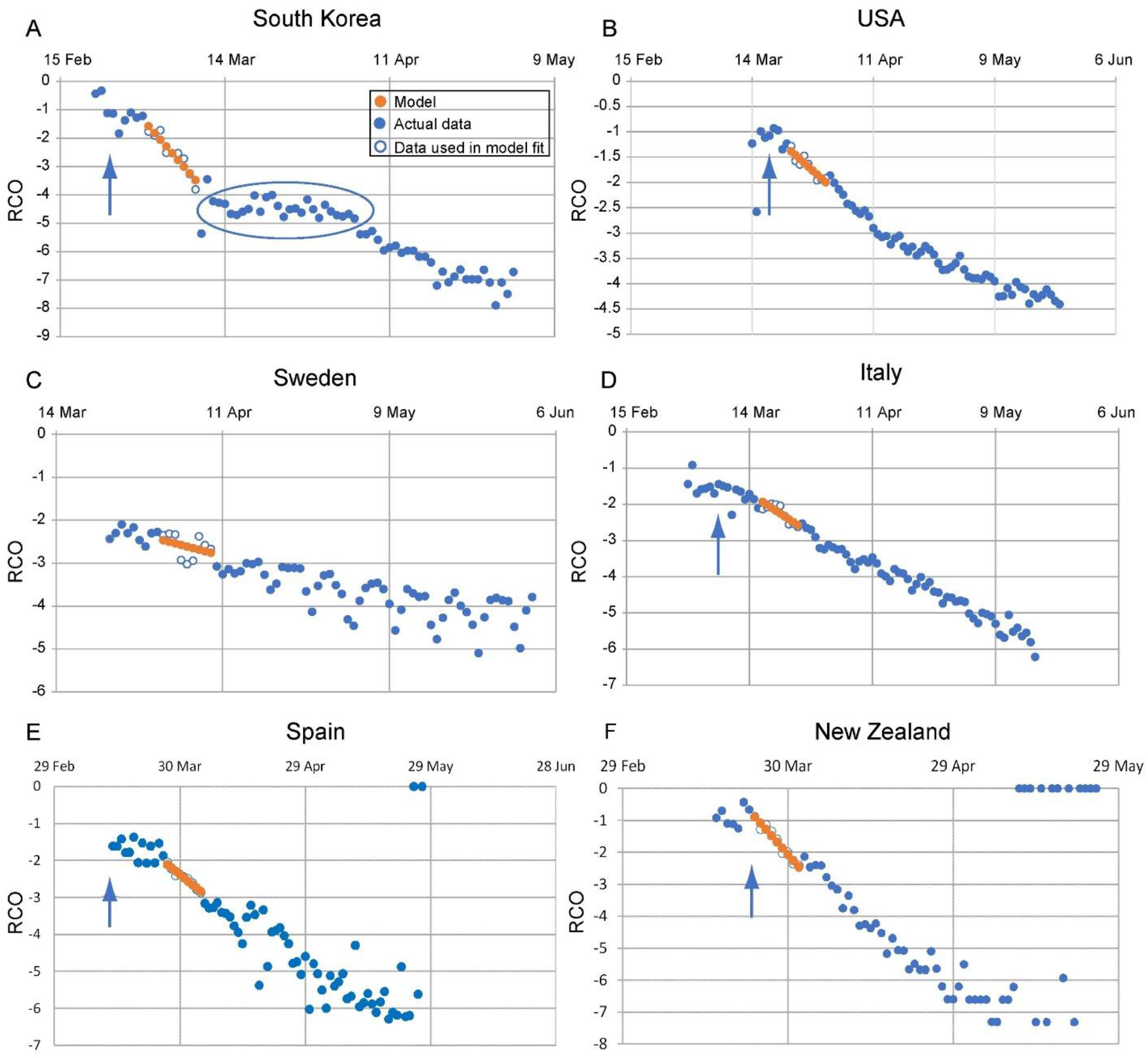
Rate of change operator (RCO) curves for COVID-19 cases in various countries. An epidemic can be described by a piecewise linear model using the RCO (Equation 42). A short segment of orange dots in each graph is a linear fit to the corresponding points (blue/white circles) in the observed data. The slopes of these dotted-line segments are the values of 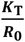as tabulated in Table2. In some countries, RCO curves changed markedly soon after the date containment measures were implemented (arrows): **A)** South Korea, February 21; (the oval highlights a departure of the observed data from the RCO slope, indicating failures in, or relaxations of, social distancing); **B)** USA, March 16; **C)** Sweden did not implement any specific containment measures, so the short line fit was begun on April 1, the date when the slope of the RCO curve first became steady. **D)** Italy, March 8; **E)** Spain, March 14; **F)** New Zealand, March 25. All dates are in 2020.

Figure 3 depicts the KMES projections of the daily total case data using the parameters from Table 2 in Equation 43. Each projection matches the actual time series of the total cases with an *R*^*2*^ >0.93 and >0.78 for 30 days and 60 days, respectively, after the first date (*t*_1_) of the date range in Table 2. Likewise, the projected values of daily <u>new</u> cases estimated from the data in Figure 3 appear in Figure 4. The *R*^*2*^ of these projections range from 0.04 (Sweden) to 0.89 (Italy); and the times projected for the peaks closely mirror the observed peak positions.

**Figure 3.**
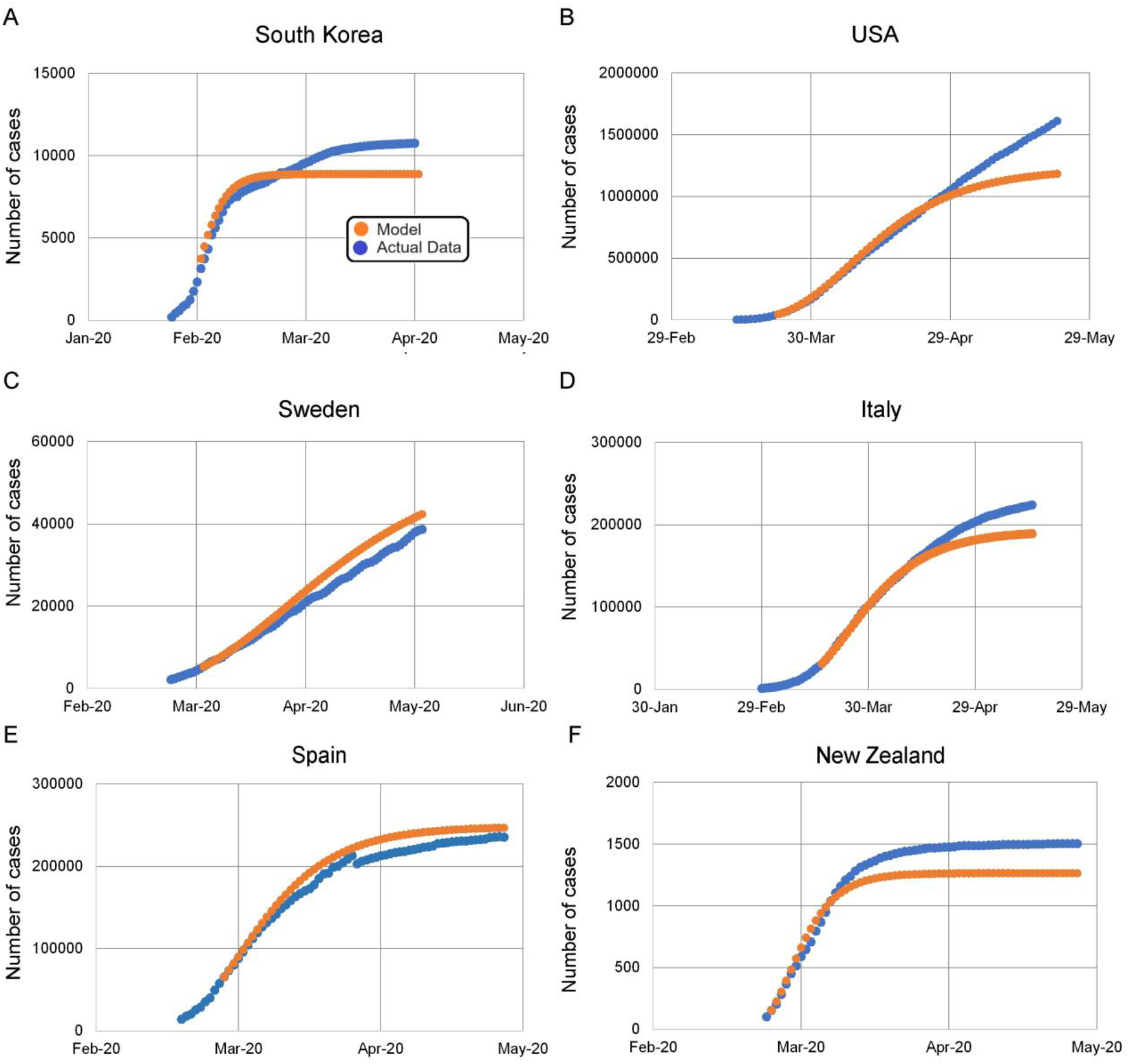
KMES model projections for daily total case counts. **A)** South Korea; **B)** USA; **C)** Sweden; **D)** Italy; **E)** Spain; and **F)** New Zealand. Dots are daily data points observed from (blue) or calculated (orange) for each country. Table 2 lists the KMES model parameters used in these plots. We found these parameter values by fitting the RCO over the date ranges listed in Table 2 and shown in Figure 2. *R*^2^ >0.93 and 0.78 for the model projections in all countries for the 30 and 60 days, respectively, after the first date in the date range shown in Table 1. The 60 day periods are: South Korea, March 1-April 30; USA, March 23-May 22; Italy, March 17–May 16; Spain, March 27-May 26; New Zealand, March 24-May 23. Sweden April 1-May 31. We explain the deviation of the model from the data in the USA, panel (**B**), after April in Supplement 2. All dates are in 2020.

**Figure 4.**
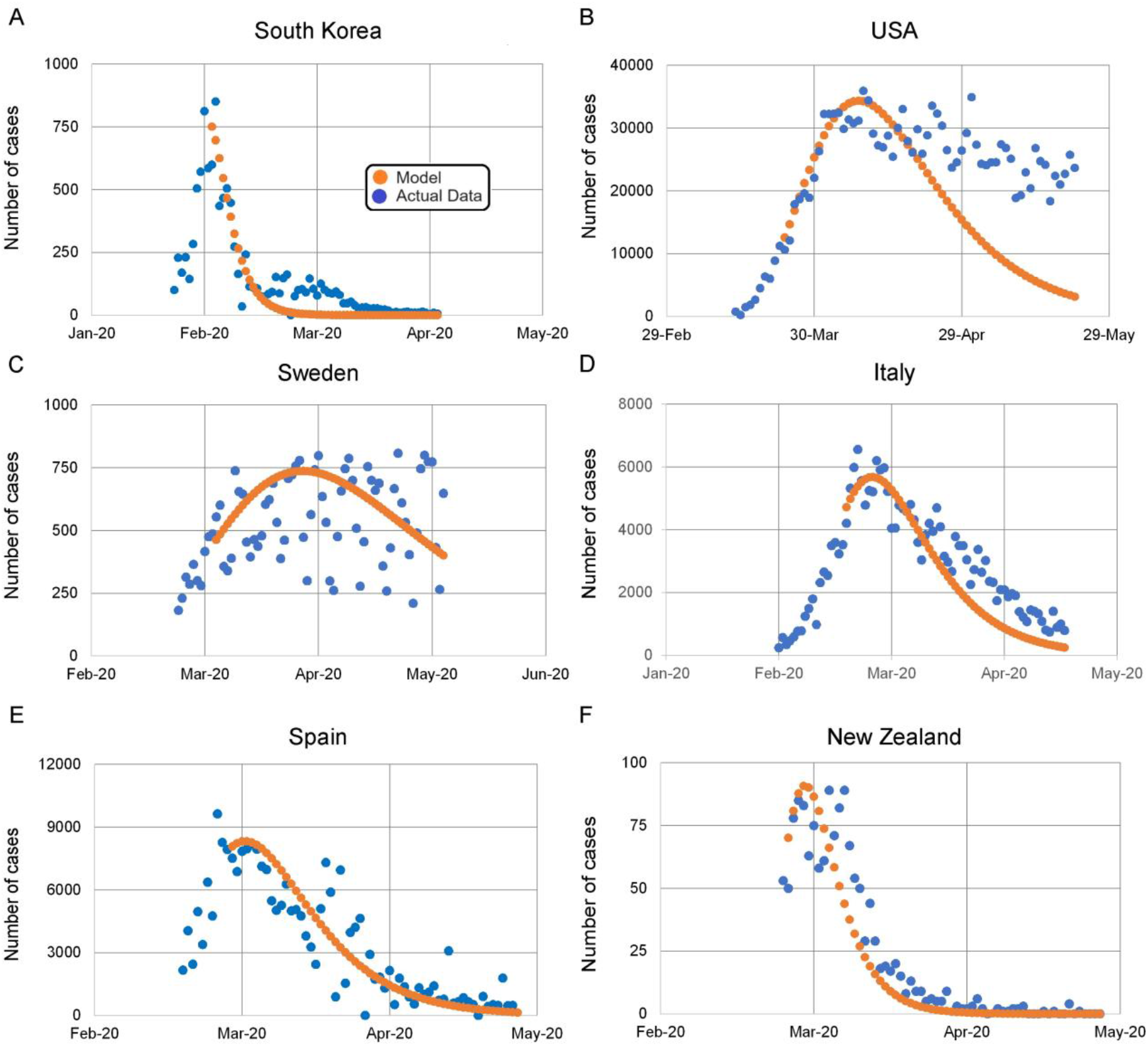
KMES model projections for number of new daily cases. **A)** South Korea, *R*^2^ = 0.86; **B)** USA, *R*^2^ = 0.32; **C)** Sweden, *R*^2^ = 0.04; **D)** Italy, *R*^2^ = 0.89; **E)** Spain, *R*^2^ = 0.81; and **F)** New Zealand, *R*^2^ = 0.85. The orange dotted line is the model in all panels. The blue dots are the daily observations from each country. The. *R*^2^ values are between the model and the data, across countries for the 60 days after the date of the first date in the date range shown in Table 2.

We emphasize that Figures 3 and 4 are not curve fits, but rather projections of the future epidemic dynamics for 4 and more than 7 weeks into the future after the date range used to estimate the values of 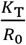. Of course, we should expect that changes in population interactions that occurred within the projection period will diminish the precision of the projections. Indeed, in Supplements 2 and 3, we discuss documented changes in South Korea and the US during these periods which explain the inaccuracies in the latter weeks noted in the figure captions. In practice, if public health officials were to use the KMES to inform their decisions, the RCO would be continuously updated, and the projections from such updates would track the changing situation in real time at, or better than, the accuracies of our 30 day projections.

### Contagiousness

Since *B*(*t, t*)*I*(0) is the time varying infectiousness of the initial infectors and depends upon contact with susceptible persons, the time course of contagiousness is calculable by eliminating the effect of the diminishing susceptible persons. Therefore, in an imaginary scenario, if we assume that *R*_0_ = 1 (i.e., the initially infected person will infect, on average, only one other person), the normalized contagiousness of the initially infected person is, This expression is solely dependent on *K*_*T*_, supporting the assumption that *K*_*T*_ is a disease parameter.

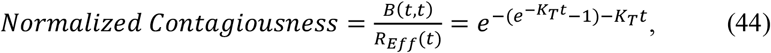

### Estimation of *K*_*T*_

To evaluate Equation 44, we must find a suitable value for *K*_*T*_. In pursuit of this goal, we first assume that, early in the epidemic, there is a relationship between *R*_0_ and the population density. Moreover, because the population of a country typically is only physically present within ∼1% of the land in a region (Ritchie and Roser 2019), we can then define an “effective area” parameter, *A*_1_(*t*), where,

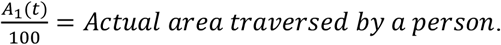

If we assume that an individual’s infectious mobility extends over an average effective area per unit of time, we can then write an expression in terms of this area and define *A*_1_(*t*) as,

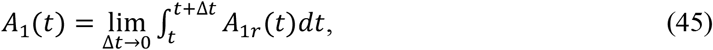

where *A*_1*r*_(*t*) equals the effective area per unit time that an individual infectiously inhabits. If we assume that changes in the value of *A*_1_(*t*) occur slowly in time, we can write a new expression for 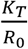,

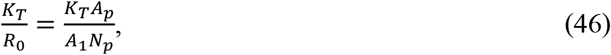

where *N*_*p*_ is the entire population of the region containing the infection, *A*_*p*_ is the area of the region, 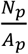 is the population density, and *A*_1_ is a constant.

Substituting Equation 46 into Equation 26 and solving for −*K*_*T*_*t*, yields,

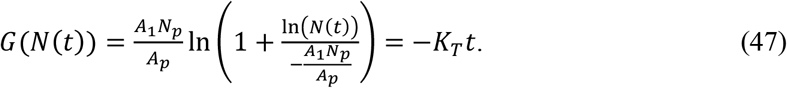

Since, other than *K*_*T*_ and *A*_1_, the quantities in Equation 47 are available from independent sources, we can use this equation to determine the values of *K*_*T*_ and *A*_1_ from early epidemic data. This procedure has two steps: first, we use an iterative process to find the value of *A*_1_ that produces the best fit of Equation 47 to the available data; and second, we evaluate *K*_*T*_ as the slope of this best fit line.

Table 3 lists the available early pandemic data for our six sample countries. Using this data, through a process of iteration, we produced Figure 5 and found that the best fit (R^2^ = 0.96) occurs when *A*_1_ = 0.48 km^2^. Consequently, by the definition of *A*_1_, the estimated <u>actual</u> area traversed by a person is 4800 m^2^; a sensible result since 4800 m^2^ = 80 m x 60 m is a plausible area for a person to traverse in a day.

**Table 3.** Initial COVID-19 pandemic data and population densities for various countries ((Roser et al 2021), case and date data; (Worldometers 2021, population density data)

| | Date first case reported | Calculation date | Days between the first case reported and the calculation date | Cases on calculation date, $N(t)$ | Population density (people/$\text{km}^2$) |
| --- | --- | --- | --- | --- | --- |
| **South Korea** | 22 Jan | 21 Feb | 30 | 204 | 527 |
| **USA** | 22 Jan | 19 Mar | 57 | 13,663 | 36 |
| **Sweden** | 1 Feb | 7 Mar | 35 | 179 | 25 |
| **Italy** | 31 Jan | 24 Feb | 24 | 229 | 206 |
| **Spain** | 1 Feb | 13 Mar | 41 | 5,232 | 94 |
| **New Zealand** | 28 Feb | 19 Mar | 20 | 28 | 18 |

All dates are in 2020.

**Figure 5.**
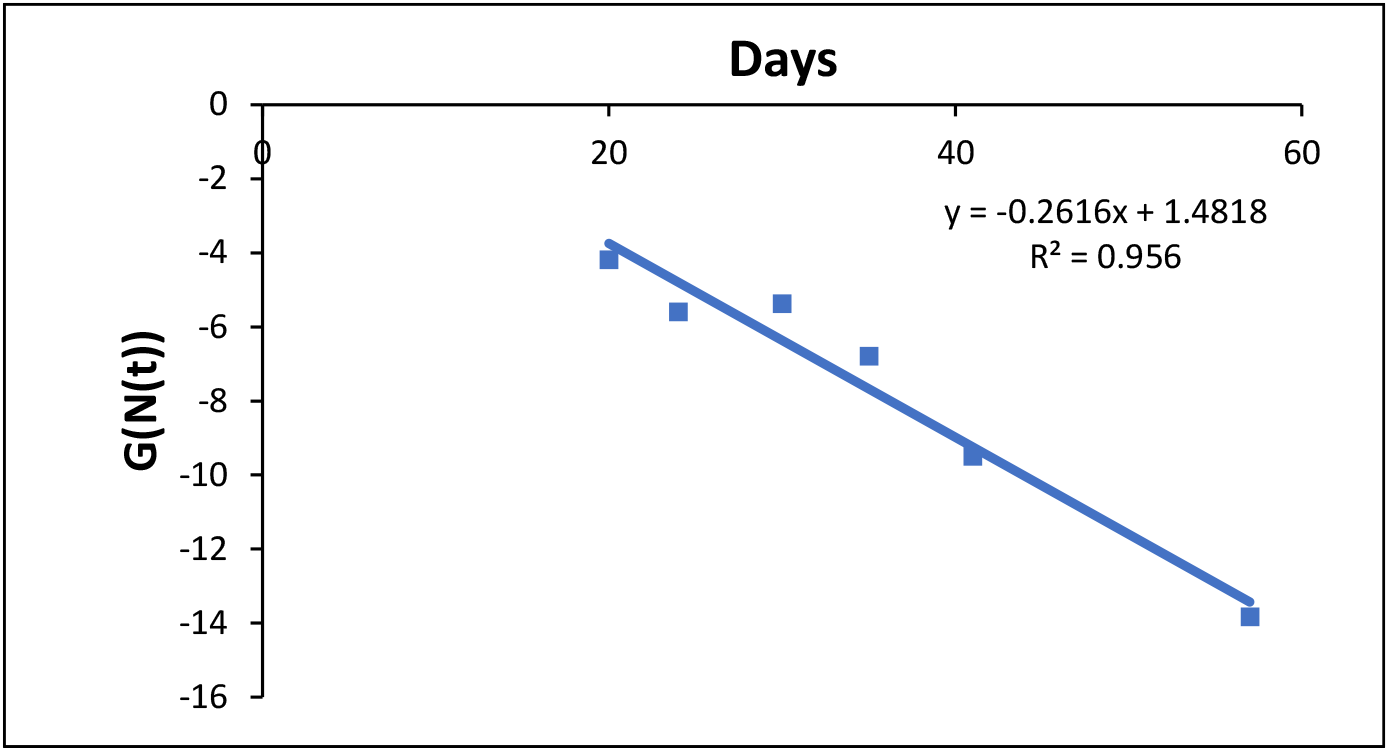
Verification that *K*_*T*_ may be the same for all countries. The data from Table 3 is plotted using Equation 47 and *A*_1_ = 0.48 *km*^2^. Each data point corresponds to a different country. The fitted value of *K*_*T*_ is the negative of the slope of the line, and *K*_*T*_ ≈ 0.26 everywhere along the line.

In Figure 5, the line has a fitted slope of 0.26, the putative value of *K*_*T*_, with a 95% confidence interval (CI) from 0.4 to 0.13. This well-fit line is strong support for both the assumption that *K*_*T*_ is a parameter of the disease, and that *K*_*T*_ and *A*_1_ were constant across the sampled countries in the initial stages of the pandemic.

Figure 6 is a plot of the contagiousness (Equation 44) created using the fitted and 95% CI values for *K*_*T*_. The solid curve in the figure suggests that an individual infected with COVID-19 becomes contagious beginning ∼5 days before the peak and declines to minimal contagiousness ∼10 days after the peak. This timing corresponds with reported values of viral shedding measured in individual patients (Puhach et al 2023, Figures 2 and 3).

**Figure 6.**
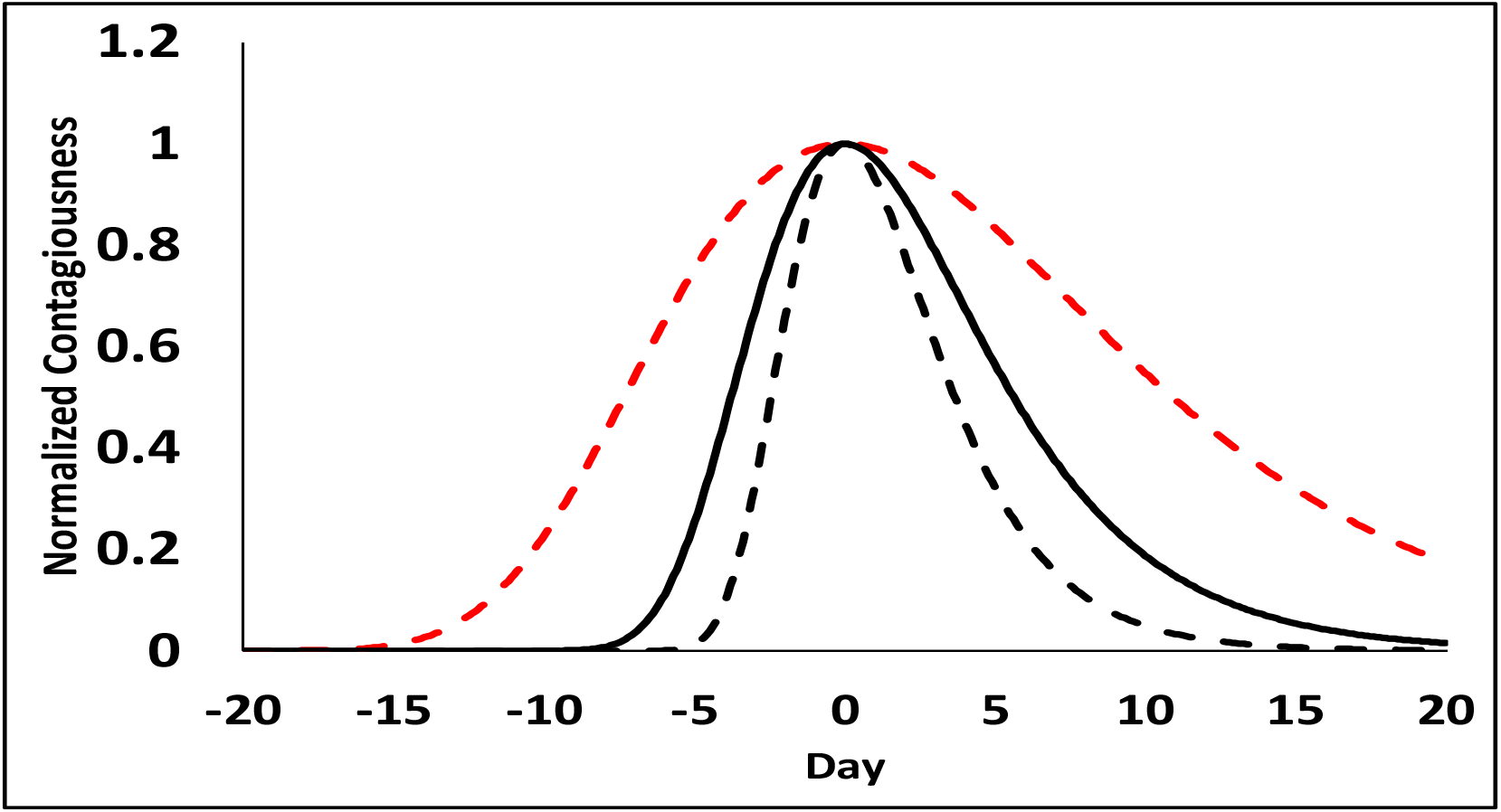
Representation of the normalized contagiousnesss of an initial infected person with one contact. The solid line was generated using the value of *K*_*T*_ = 0.26 in Equation 44; the black dashed line *K*_*T*_ = 0.40; and the red dashed line, *K*_*T*_ = 0.13; the fitted and the upper and lower 95% confidence levels, respectively, for *K*_*T*_. The contagiousess begins before time zero because the initial individual was infected before the start of the epidemic.

## Discussion

In response to Diekmann’s call for action, the “wisdom” we found in the Kermack and McKendrick integro-differential equations appears substantial. Foremost, we offer that the KMES obviates the need to use any approximation to the integro-differential equations because it is an exact solution and its two-parameter construction is even simpler than, for instance, the three-parameter basic SIR model approximation. Secondly, the KMES has an intuitive probabilistic form which appears to enable accurate projections of reported pandemic data dynamics and offers a pathway to new analytical expressions for characterizing and managing epidemics.

The analysis hinges on the N-group perspective, an equivalent regrouping of the parameters in K&M’s integro-differential equations. Using *R*_0_ as the point of reference, this perspective employs K&M’s original concept of capturing epidemic dynamics by tracking and summing the creation and recovery of infections, but, in contrast to K&M’s approach, it does so in a manner that produces resolvable expressions.

These expressions subsume the susceptible total into *K*_*T*_; a favorable formulation which derives from the insight that a contagious individual is infectious proportionate to their contact with the susceptible people that they <u>can</u> infect. Inspired by the N-group perspective, this use of *K*_*T*_ circumvents the assumption of a well-mixed population and emphasizes that the rate of new infections must be directly dependent on the contemporaneous infectiousness. In tandem, the N-group perspective enables us to easily track the total infectiousness of the groups and tie this total to the quantity of ever-infected persons and their recovery.

Aiding the solution derivation, the utilization of the weighted averages, 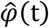 and 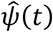, eliminates the variable *θ* and creates simpler, time-dependent differential equations. Taken altogether, these insights do not change the character of K&M’s equations; rather, by adopting a more practical method to sum up the population variables, enabled by a change in perspective, they make the equations solvable.

We underscore that the closed form of the KMES rests squarely on the assumption that *K*_*T*_ and *R*_0_ are constant for the duration of the epidemic. This assumption, though plausible, is an important limitation on the utility of the KMES. For example, in Supplements 2 and 3 we hypothesize that any large divergence between the KMES projections and the measured data in Figure 3 may be due to fluctuations in the population interactions (and thus *R*_0_) that drove the outcomes in the early portions of the COVID-19 pandemic. A deeper understanding of the nature of the cause-and-effect relationship between *R*_0_ and the true population interactions could be gained by using an independent measure of population behavior in our solution as a proxy for *R*_0_, but this can only be studied if we have a solution wherein *K*_*T*_ and *R*_0_ are not required to be constant.

As a second example, a disease with a latent period could manifest itself by affecting the value of *K*_*T*_, potentially requiring it to vary in time. We did not explore the analytical consequences of this case; however, we could develop the solution further by first separating the population of latent infections from the contagious infections within *I*(*t*). Then, by exploring the effect of the latent period on the solution parameters, we could determine whether the latent period can be found early in an epidemic and/or ignored once the epidemic has fully developed. Both would be useful.

A future publication will address the complexities of modelling the variation in social response during epidemics with the presentation of an explicit solution to the K&M equations for the circumstance in which *K*_*T*_ and *R*_0_ do vary with time. However, due to the complexity of allowing temporal variation, we have restricted the current manuscript to the insight and opportunity that derives from a closed form solution.

The hybrid path, combining network concepts and deterministic equations, that we used to derive the KMES, hints that herein may lie the potential to develop more sophisticated hybrids. Further exploration of the means to specify *R*_0_ within types of networks representing measured physical interactions between people may well lead to more accurate forecasts of epidemic progressions. Such extensions could be useful when applied to influenza epidemic data and data from a myriad of other diseases, including RSV and the next SARS virus redux. Hybrid approaches such as these could also reduce the computational burden associated with models that employ high numbers of network nodes by allowing the calculation of results for portions of the network using analytical expressions rather than simulations.

Our analysis can be further improved by systematic address of the enormous amount of case data now available from the COVID-19 pandemic. This added analysis should refine the process required to estimate the key parameter, *K*_*T*_, including variations in time, mutations of the infectious agent and local genetic variations in the affected population. In the inevitable future epidemics, this could substantially improve the analyses aimed at establishing the actions people and governments need to take to achieve target values of *R*_0_.

These potential improvements notwithstanding, the closed form solution of the KMES supports and explains the findings of other researchers. For example, Equation 26 is a Gompertz equation, and as Onishi et al (2021) and Pelinovsky et al (2022) highlighted, the COVID-19 epidemic time course in many countries was well fit by a Gompertz model. These authors did not offer a basic epidemics principles argument for this finding. The KMES does.

Furthermore, the KMES points to a simple method to assess whether an outbreak is occurring. Embodied in *R*_*eff*_, this offers an additional quantitative tool to guide public health officials’ decision making. We illustrate this in Supplement 2.

Two conclusions, which are in contradistinction to conventional epidemic concepts, derive from the KMES. The first, an expression for the time to the peak number of new cases, Equation 40, shows that if people interact less often and decrease *R*_0_, the peak number of infections will be much lower, and will occur much earlier; the antipode of this relationship to *β* in SIR models and the resolution of the paradox presented in the introduction. In support of our conclusion, we submit that the KMES correctly and intuitively projected much lower and earlier peaks in the two countries, New Zealand and South Korea, which imposed strong containment strategies compared to the other four sample countries (Figure 4). Significantly, Sweden and Italy, two countries similar in size to New Zealand and South Korea with less stringent containment strategies, had higher and later peaks.

The second heterodoxic conclusion, the final size expression in Equation 41, asserts that the K&M equations allow for the entire population to be infected, and lies in opposition to the conventional expression for herd immunity. We rest our arguments on the mathematical proof that the KMES solves the K&M equations, and that the final size expression derives from the KMES.

As a last point, and as added insight into the KMES, we highlight that

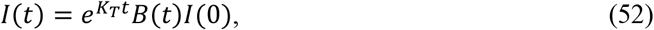

is an alternative form of the solution for *I*(*t*). This equation states that the input of infections, *I*(0), equates to the time varying output of infectiousness, *I*(*t*), when multiplied by a step response function,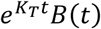. Therefore, in addition to our probabilistic interpretations, an epidemic can be cast as a dynamic system responding to a step input amplified by a function dependent on the disease and population behavior; an obvious-in-retrospect mathematical characterization.

## Data Availability

All data produced in the present study are available upon reasonable request to the authors

## Author contributions

T. Duclos and T. Reichert: Conceptualization, Writing – Original draft preparation, Writing – Review & Editing. T. Duclos: Methodology, Validation. T. Reichert: Data Investigation.

## Abbreviations used in this text

KMES: Kermack McKendrick integro-differential Equation Solution
K&M: Kermack and McKendrick
RCO: Rate of Change Operator
SIR: Susceptible–Infectious–Recovered
CI: Confidence Interval

## Supplement 1: *I*(*t*) as a Function of 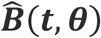

We know from K&M (1927) that 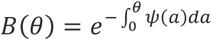, and from Equation 2,

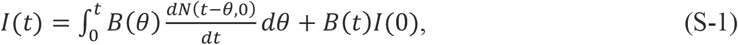

Our goal in this supplement is to derive an equivalent equation for *I*(*t*) in terms of the infection probability function 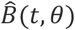, which is based on 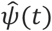, the weighted average of *ψ*(*θ*).

We begin by noting that since 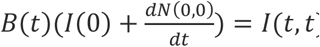, Equation S-1 reduces to,

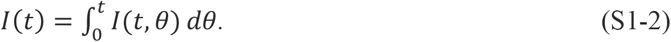

From Equation 3, Equation S1-2, and the definition of 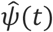, we can write,

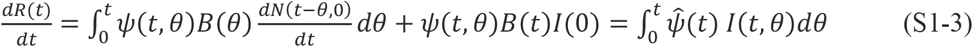

The derivation begins with the change in the total infectiousness,

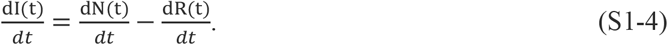

Equation S1-4 can be discretized as,

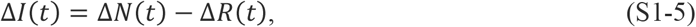

and rewritten using the discrete forms of Equations S1-2 and S1-3, with the equality Δ*t* = Δ*θ*, as,

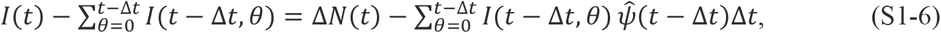

Then,

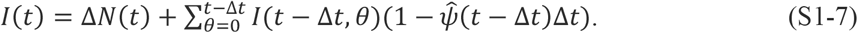

We begin a useful sequence by expanding the right-hand side of Equation S1-7 by one term,

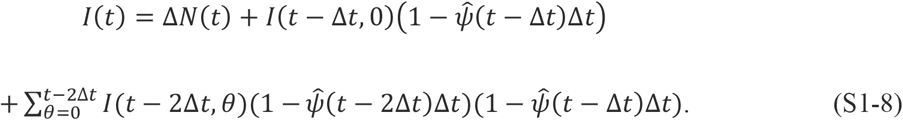

Continually writing out terms on the righthand side, and since Δ*N*(*t*) = *I*(*t*, 0), the new infections at time, t, we generate the following sequence,

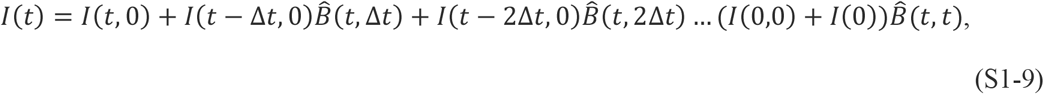

where,

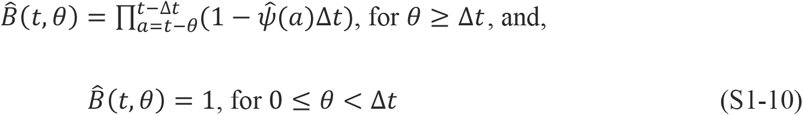

Taking the limit of S-10 as Δ*t* goes to zero, we find that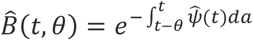.

This important result shows that the integral limits in 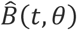 must be the time interval from *t* − θ to t, a time of length *θ*; the same time interval covered in K&M’s formula for *B*(*θ*). Furthermore, since 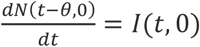, we deduce from Equations S1-1, S1-9 and S1-10,

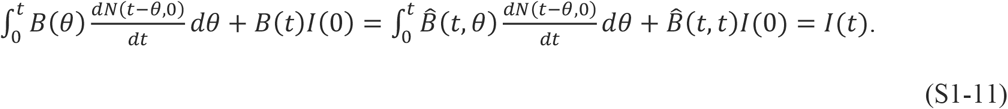

## Supplement 2. Controlling epidemics early

The relationships derived from the KMES in Section 1 illustrate that strong and early intervention is not only critical for control at any time, but especially early in the epidemic and at every genetic change in the disease agent. For instance, Equation 41 quantifies that the final number of individuals infected in an epidemic depends *exponentially* on the number of people with whom each person interacts.

Real-world country data exemplifies these consequences. Both South Korea and New Zealand enacted strong and early interventions compared to other countries (Campbell, C 2020; Field, A 2020), as reflected by their higher 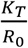 values (Table 2). These interventions led to earlier peaks in new cases and to far fewer total cases than in other countries (Figures 3 and 4) in the first months of the pandemic. The peak number of new cases in both South Korea and New Zealand was 90–99% lower than in other countries, a validation of the statement in the KMES that strong interventions lead to *exponentially* better outcomes.

In the USA, interventions started on March 16 began to have an effect around March 23, 2020 (Figure 2B), and the number of new cases on March 23, 2020 (Roser et al 2021) was 46,136. Using the values of ln (*F* _*i*_ (*t*_1_)*K* _*T*_(t)) and 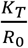 in Table 2, Equation 41 predicts that had these remained constants, the ultimate number of cases would have been approximately 1.22 million. On the other hand, if public officials and the population had attained the same intervention, on March 10, when there were 59 times fewer (782) cases (Roser et al 2021), then the model predicts that the number of cases would also have been 59 times lower, or 20,725. Thus, earlier action could have reduced the number of projected cases by more than 98%.

Of course, the projected estimate of 1.22 million total USA cases would only have occurred if the populace had sustained the effectiveness of the interventions launched on March 16. Unfortunately, a marked reduction in effective interventions occurred in parts of the USA in mid-April, well before the official reopening of the economy (Elassar 2020). This caused a second surge in new cases in late April and is why the observed data and the KMES predictions diverge in Figures 3B and 4B.

As shown in Section 1, Equation 39 furnishes an estimate of the time to the peak of new cases, *t*_peak_. By using the data from Table 2 in Equation 39, we predict that the peak in new cases in the USA would have occurred near March 24 if the intervention had begun on March 10.

Instead, a 6-day delay in intervention shifted the initial peak to April 11, with a much higher value (Figure 4B).

Epidemic acceleration, obtained by differentiating Equation 36, and then applying the definition of the RCO,

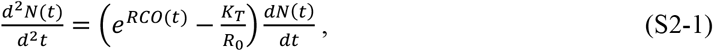

is calculable at any time. While its value depends on the social containment actions in effect, what is perhaps less apparent is that two countries with identical new case counts on a given day with different accelerations, will exhibit different dynamics immediately after that day.

For example, South Korea and New Zealand (Figure 4A and F) had similar new case counts (204 cases in South Korea on February 21, and 205 in New Zealand on March 25) when each imposed similar containment measures (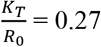in South Korea and 0.20 in New Zealand; see Table 2). However, as evidenced by its higher RCO at the time of intervention, South Korea had a higher acceleration on February 21 compared to New Zealand on March 25. Consequently, the later number of cases in South Korea was higher than in New Zealand (Figure 3A and F).

Equation 20 clearly illustrates these lessons. Strengthening social distancing lowers *R*_*eff*_(*t*) by reducing *R*_0_ and, thereby, the epidemic slows. Earlier and stronger interventions in countries with indigenously elevated levels of social interaction, are necessary to stop an epidemic in the first stages. Reopening, enacted too early, reignites an epidemic, considerably increasing the number of cases. The immense difference in the size of the effects, driven by a few days of delay, derives from the doubly exponential nature of the underlying relationships.

## Supplement 3. Controlling and Ending an Ongoing Epidemic

If the situation warrants, the time needed to reach a desired reduction in 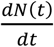, at a future time, *t* + *t*_*target*_, can be calculated for a given level of social interaction by first defining the desired fractional reduction in cases as *D*_*tf*_,

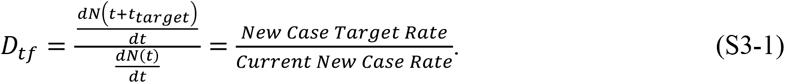

Using Equation 36, the following expression emerges from Equation S3-1,

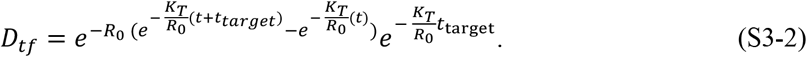

If *t* ≫ *t*_*target*_, then 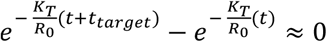, and also,

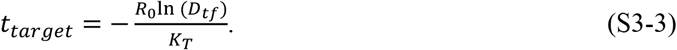

Equation S3-3 quantitates the number of days, *t*_*target*_, that a level of social containment, *R*_0_, must continue to reach a desired reduction of the current daily cases.

Equation S3-3 projects the number of days the populace must sustain a given level of intervention, 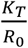, to reduce the new daily cases by a target fraction, *D* _*tf*_. Thus, a country targeting a 90% reduction of new cases per day (e.g., from 50,000 to 5,000 cases per day, *D*_*tf*_ = 0.1), can attain its target in about 12 days by imposing a containment level of 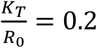.

As an example of this scenario, the South Korea and New Zealand data show that both countries achieved a value of 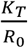 close to 0.2 for the time necessary to produce a 90% reduction. In agreement with the KMES, it took 13 days for South Korea (March 3–16) and 15 days for New Zealand (April 2–15) to reduce their new cases by 90% between the dates shown.

Returning to the planning example, after achieving the first 90% reduction, a reasonable next step might be to relax social containment to a level that allows the economy to still be viable, while preventing the epidemic from erupting again. We can again find the level of 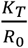 necessary to achieve a chosen target using Equation S3-3. If an additional 90% reduction in new cases per day is desired, and a period of 90 days is tolerable to achieve that reduction, then affected population must attain and maintain a new interaction level where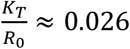. This equates to a 90-day period during which each person can on average infectiously contact ten people. Note that this is three times *less* stringent than the original USA shutdown level in April (Table 2). Thus, with a well-planned approach, a country can reduce its new daily cases by 99% in approximately 100 days, control the epidemic, and simultaneously maintain economic viability.

If even 0.026 is too restrictive, we can choose a still lower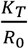, but it must be large enough to avoid a new outbreak. A lower bound for the new value of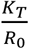, high enough to prevent an outbreak, can be found by setting the left-hand side of Equation S2-1 to zero, then solving for 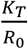.

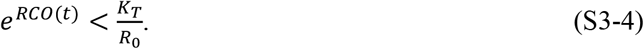

We can easily monitor the progress of interventions using the RCO (Equation 42), as the RCO curve for South Korea illustrates (Figure 2A). Had this country maintained the initial level of distancing measures, the data would have followed the initial slope. However, the actual data departed from the slope, heralding that failures in (or relaxation of) social distancing, occurred during the indicated period (Campbell 2020) (circled data, Figure 2A). Because it summarizes epidemic dynamics, we can use the RCO to continuously determine the effectiveness of implemented measures and whether they need adjustment.

## Supplement 3.1 Outbreaks

We can see from Equation 42 that if the social interactions decline (lower *R*_0_), the slope of the RCO curve steepens, and, conversely, if the interactions increase, the slope lessens.

Therefore, if the value of *K*_*T*_ does not change due to a change in the disease transmissibility, the RCO slope is a metric for monitoring population interactions. It is also clear that, under the assumptions used to develop the KMES, 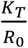 must always be greater than zero, and the RCO slope can never become positive. However, this only holds under three conditions: 1) immunity persists, 2) no new infections are introduced from outside the area, 3) any change, Δ*R*_0_(*t*), due to new susceptible people entering existing N-groups or infectious people entering previously unaffected population areas must be an order of magnitude smaller than the new infections, *K*_*T*_(*t*)Δ*t*. We call the latter two conditions the assumption that the epidemic is contiguous.

If new infections enter a portion of the population consisting only of susceptible people, then the assumption of contiguousness does not hold. This is a common situation when infected people travel from an infected area to a previously uninfected area and cause an outbreak.

If assumptions 2 and 3 no longer hold, to predict the number of cases in an epidemic affected by an outbreak, we must modify Equation 26. If when *t* = 0, *N*(0) = 1; and introducing the notation *R*_0*x*_ where *x* denotes the number (order in time) of the outbreak, Equation 26 becomes:

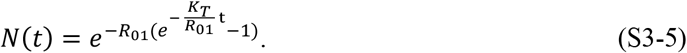

If a new outbreak occurs in a previously unaffected area of a country, then we modify Equation S3-5 as follows:

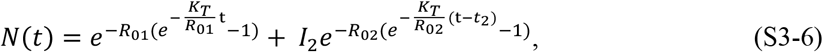

where *I*_2_ is the number of infectious people who started the new outbreak, *R*_02_ is the replication number in the new outbreak area, and *t*_2_ is the time the new outbreak occurs. Although we have assumed that *K*_*T*_ remains the same throughout this illustration, if *K*_*T*_ changes in a subset of the population, then the notation, *K*_*Tx*_, can track the populations with the new transmissibility.

Equation S3-6 written in a general form is,

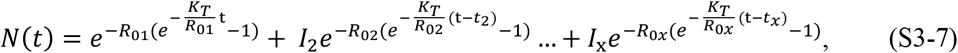

where *x* denotes the outbreak number and t > *t*_2_ > *t*_3_ > ⋯ > *t*_*x*_. For each outbreak *t*_*x*_, *R*_0*x*_, and *I*_*x*_ need to be found independently.

While an epidemic is underway a positive slope detected in an RCO curve indicates that an outbreak has occurred, and that policy makers must take immediate action, within days, to strengthen intervention measures if they are to prevent the outbreak from overwhelming prior progress in controlling the epidemic.

## References

Brauer, F. Compartmental Models in Epidemiology. In: Brauer, F., van den Driessche, P., Wu, J. (eds) Mathematical Epidemiology. Lecture Notes in Mathematics, 2008, vol 1945. Springer, Berlin, Heidelberg. 10.1007/978-3-540-78911-6_2

Breda D, Diekmann O, de Graaf W F, Pugliese A, Vermiglio R On the formulation of epidemic models (an appraisal of Kermack and McKendrick). Journal of Biological Dynamics, 2021, 6:sup2, 103–117, DOI: 10.1080/17513758.2012.716454

Campbell C. South Korea’s health minister on how his country is beating coronavirus without a lockdown. TIME. https://time.com/5830594/south-korea-covid19-coronavirus/ (2020). Published April 30, 2020. Accessed February 9, 2021.

Di Lauro F, Berthouze L, Dorey MD, Miller JC, Kiss IZ. The Impact of Contact Structure and Mixing on Control Measures and Disease-Induced Herd Immunity in Epidemic Models: A Mean-Field Model Perspective, Bulletin of Mathematical Biology (2021) 83:117 10.1007/s11538-021-00947-8

Diekmann O, European Communications in Mathematical and Theoretical Biology 2022 No. 24, pgs. 7–9

Diekmann O, De Jong MCM, Metz JAJ, A Deterministic Epidemic Model Taking Account of Repeated Contacts between the Same Individuals Journal of Applied Probability, Vol. 35, No. 2. (June 1998), pp. 448–462

Diekmann O, Heesterbeek H, and Britton T, Mathematical Tools for Understanding Infectious Disease Dynamics, Princeton University Press (2013), ISBN 978-0-691-15539-5

Diekmann O, Othmer H G, Planque R, Bootsma, M C. The discrete-time Kermack– McKendrick model: A versatile and computationally attractive framework for modeling epidemics. PNAS 2021 Vol. 118 No. 39 e2106332118. 10.1073/pnas.2106332118

Elassar A, This is where each state is during its phased reopening. CNN. https://edition.cnn.com/interactive/2020/us/states-reopen-coronavirus-trnd. Published May 27, 2020. Accessed February 9, 2021.

Field A, New Zealand isn’t just flattening the curve. It’s squashing it. The Washington Post. https://www.washingtonpost.com/world/asia_pacific/new-zealand-isnt-just-flattening-the-curve-its-squashing-it/2020/04/07/6cab3a4a-7822-11ea-a311-adb1344719a9_story.html. Published April 7, 2020. Accessed February 9, 2021.

Hethcote HW. The Mathematics of Infectious Diseases. SIAM Review December 2000, Vol. 42 No. 4, 599–653

Kroger M, Schlickheiser R, Journal of Physics A: Mathematical and Thoeretical, 53, 2021, 10.1088/1751-8121/abc65d

Kermack WO, McKendrick AG. A contribution to the mathematical theory of epidemics, Proc R Soc Lond A. 1927, 115(772):700–721. 10.1098/rspa.1927.0118

Newman, MEJ, Spread of epidemic disease on networks, PHYSICAL REVIEW E 66, 016128, 2002

Ohnishi A, Namekawa Y, Fukui T. Universality in COVID-19 spread in view of the Gompertz function. Prog. Theor. Exp. Phys. 2020, 12:123J01. 10.1093/ptep/ptaa148

Pelinovsky E, Kokoulina M, Epifanova A, Kurkin A, Kurkina O, Tang M, Macau E, Kirillin M, Gompertz model in COVID-19 spreading simulation, Chaos, Solitons & Fractals, Volume 154, 2022, 111699, ISSN 0960-0779, 10.1016/j.chaos.2021.111699

Puhach, O., Meyer, B. & Eckerle, I. SARS-CoV-2 Viral Load and Shedding Kinetics. Nat Rev Microbiol 21, 147–161 (2023). 10.1038/s41579-022-00822-w

Ritchie H, Roser M. Land use. Our world in data. https://ourworldindata.org/land-use Published September 2019. Accessed February 9, 2021.

Roser M, Ritchie H, Ortiz-Ospina E, et al. Coronavirus pandemic (COVID-19). Our World in Data. https://ourworldindata.org/coronavirus. Published December 2, 2021. Accessed December 2, 2021.

Worldometers. https://.worldometers.info. Published August 21, 2021. Accessed August 21, 2021

Youssef M, Scoglio C, An individual-based approach to SIR epidemics in contact networks, Journal of Theoretical Biology, Volume 283, Issue 1, 21 August 2011, Pages 136–144, 10.1016/j.jtbi.2011.05.029

Zonta F, Levitt M, Virus spread on a scale-free network reproduces the Gompertz growth observed in isolated COVID-19 outbreaks, Advances in Biological Regulation, Volume 86, 2022, 100915, ISSN 2212-4926, 10.1016/j.jbior.2022.100915

